# Unravelling the transcriptome of the human tuberculosis lesion and its clinical implications

**DOI:** 10.1101/2022.03.31.22273171

**Authors:** Kaori L. Fonseca, Juan José Lozano, Albert Despuig, Dominic Habgood-Coote, Julia Sidorova, Lilibeth Arias, Álvaro Del Río-Álvarez, Juan Carrillo-Reixach, Aaron Goff, Leticia Muraro Wildner, Shota Gogishvili, Keti Nikolaishvili, Natalia Shubladze, Zaza Avaliani, Pere-Joan Cardona, Federico Martinón-Torres, Antonio Salas, Alberto Gómez-Carballa, Carolina Armengol, Simon J Waddell, Myrsini Kaforou, Anne O’Garra, Sergo Vashakidze, Cristina Vilaplana

## Abstract

The granuloma is a complex structure, contributing to the overall spectrum of tuberculosis (TB). We characterised 44 fresh human pulmonary TB lesion samples from 13 patients (drug-sensitive and multi-drug resistant TB) undergoing therapeutic surgery using RNA-Sequencing. We confirmed a clear separation between the granuloma and adjacent non-lesional tissue, with the granuloma samples consistently displaying increased inflammatory profile despite heterogeneity. Using weighted correlation network analysis, we identified 17 transcriptional modules associated with granulomata and demonstrated a gradient of immune-related transcript abundance according to the granuloma’s spatial organization. Furthermore, we associated the modular transcriptional signature of the TB granuloma with clinical surrogates of treatment efficacy and TB severity. We show that in patients with severe disease, the IFN/cytokine signalling and neutrophil degranulation modules were overabundant, while tissue organization and metabolism modules were under-represented. Our findings provide evidence of a relationship between clinical parameters, treatment response and immune signatures at the infection site.

---

Tuberculosis (TB) is an infectious disease caused by *Mycobacterium tuberculosis* (*Mtb*), and a major cause of ill-health and mortality worldwide^1^. Globally, TB chemotherapy is successful in 85% of drug-sensitive (DS) TB cases^2^. Nevertheless, there is a fraction of patients who will fail treatment and are therefore prone to disease relapse and death, especially in multi drug-resistant (MDR) TB cases^3^. The formation of granulomas is a hallmark of TB and is crucial for containing and controlling the spread of *Mtb* within the host^4^. The existing literature has demonstrated a high degree of heterogeneity in TB granulomatous lesions^5^. Animal studies involving macaques have provided valuable information on granuloma nature and evolution, showing high diversity even within the same host with different grades of bacteria clearance^6^. Moreover, this diversity is observed over the course of infection^7^. These preclinical studies are key to understanding how TB lesions evolve, as human studies cannot provide this information unless using surrogates, such as 18-F-fluorodeoxyglucose positron emission tomography-computed tomography (18F-FDG-PET-CT), as demonstrated by Malherbe et al. when correlating images of TB patients obtained using this method with bacillary load^8,9^.

The development of lung cavitary lesions from a granuloma is crucial in TB pathogenesis and is linked to increased transmission and poor outcomes^10^. Human TB lesion lung biopsies are limited and scarce^11^, and host factors that drive cavitary lesion formation or trigger or reflect poor clinical outcomes remain unknown. The resection of human pulmonary lesions during therapeutic surgery or autopsies has revealed insights into TB lesion architecture, and immunopathology at the site of disease, which may contribute to the emergence of MDR *Mtb* populations^12,13^. Moreover, mycobacterial culture from resected granuloma tissue demonstrated that a subset of patients still harboured live *Mtb* bacilli despite showing preoperative microbiological clearance in sputa in both, DS- and MDR-TB patients^12,14^.

Subbian et al demonstrated molecular correlation of immune responses to the heterogeneity of granuloma samples from four MDR-TB patients, diversity that the authors linked to lesion maturation^15^. Marakalala et al.^16^ suggested that the response to *Mtb* might be shaped by the anatomical localization within the granuloma^16^. They demonstrated a pro-inflammatory center and an anti-inflammatory surrounding tissue by mass spectrometry and lipid quantification. These authors worked with different types of granulomata from six MDR-TB patients and highlighted the heterogenicity of the lesions^16^. Dheda et al. were the first authors to characterize the transcriptional response at anatomically different locations within the granulomas of 14 MDR-TB patients^11^. They showed the cavity wall as the main source of pro-inflammatory activity compared to the lesion centre. However, none of these human studies included DS-TB patients. Finally, a recent study constructed a spatial cell atlas using 6 patients’ samples (two DS-TB and one MDR-TB patient undergoing surgery, and three autopsies) to map granuloma structure and composition and contrast it with the peripheral immune responses^17^.

To our knowledge, no published TB studies have correlated their results with the clinical characteristics of the patients. Based on the hypothesis that characterising human TB lesions at the transcriptomic level could help us to understand the heterogeneity of granulomas and TB pathogenesis in relation to disease presentation, we determined the modular transcriptome signatures of human TB pulmonary lesions from DS- and MDR-TB patients who underwent surgery. We also investigated their link to clinical and microbiological surrogates of TB severity and treatment response (Fig. 1).

**Figure 1.**
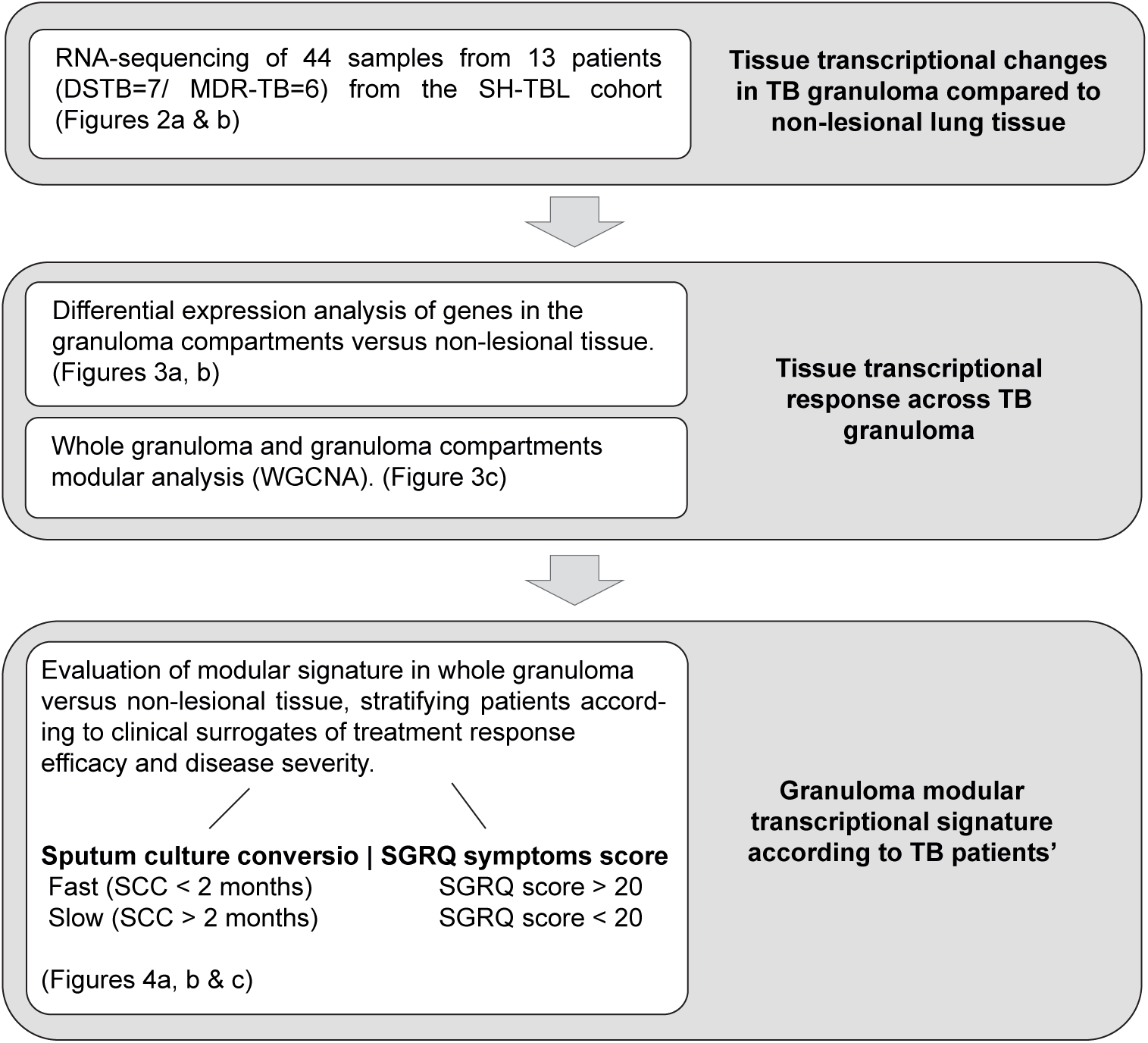
Overall study plan. Overview of the analysis undertaken in the study. Figures associated with each objective are stated.

## RESULTS

### The human TB granuloma signature shows a distinct and heterogeneous transcriptional profile as compared with non-lesional lung tissue

In our study, outlined in Fig. 1, we collected 48 samples from 14 patients and analysed 44 paired samples from 13 patients (7 DS-TB and 6 MDR-TB) to evaluate the human TB lung granuloma transcriptomic changes by RNA sequencing. We analysed total RNA from three different sections: Central Lesion (C; *n=6*), Internal Wall (I; *n=12*) and External Wall (E; *n=13*) collected from each patient’s granuloma biopsy. Fewer C-samples could be analysed compared to I and E, due to poorer RNA recovery. Additionally, surrounding non-lesional (NL) tissue from the involved lung was collected as a comparator (*n=13*) (Fig. 2a). Patients were matched according to their sex and *Mtb* drug-sensitivity classification to avoid potential confounding factors (Supplementary Table 1). Moreover, clinical and demographic data and, resected TB lesion characteristics of each participant were assessed at the time of surgery (Supplementary Tables 1 and 2).

**Figure 2.**
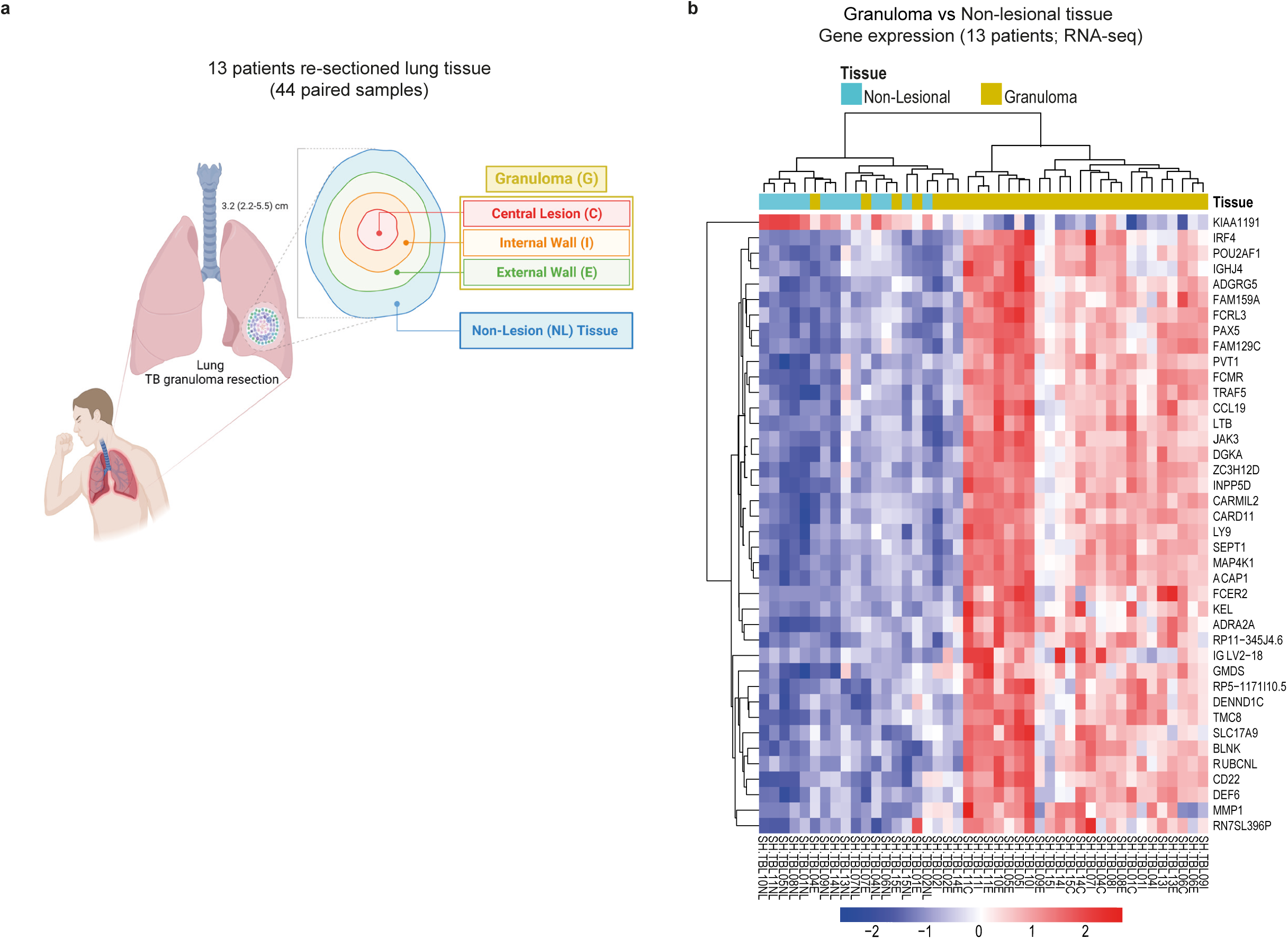
The human TB granuloma signature shows a distinct and heterogeneous transcriptional profile as compared with non-lesional lung tissue. Granuloma samples were collected from each patient included in the SH-TBL cohort: central lesion (C), internal wall (I) and external wall (E) and, altogether, samples from each patient represent the human TB granuloma (G). An additional sample from surrounding non-lesional lung tissue (NL) was also collected from the same patient as control (**Panel a**). 44 samples from 13 patients (7 DS-TB and 6 MDR/XDR-TB) were RNA sequenced to evaluate the human TB lung granuloma transcriptomic changes. A set of 4,630 DGES was identified after comparing the human TB granuloma counts with NL lung tissue expression, using DESeq2 with adjusted p<0.05. **Panel b** heatmap depicts the top 40 DEGs ranked by the adjusted p-value comparing the human TB granuloma versus NL lung tissue expression profiles. The intensity of each colour denotes the standardized ratio between each value and the average expression of each gene across all samples. Red pixels correspond to an increased abundance of mRNA in the indicated sample, whereas blue pixels indicate decreased mRNA levels.

We found a total of 4,630 significantly differentially expressed genes (DEGs), using DESeq2 with adjusted *p*≤0.05 (Extended data Fig. 1a and Supplementary Data Set 1). Of these, 2,496 genes were over-expressed in lung granuloma tissues, whereas 2,134 were under-expressed, as compared to NL lung tissue (Extended data Fig. 1a). The top 40 ranked DEGs clearly separated granuloma samples from NL lung samples (Fig. 2b), showing distinct transcriptional profiles for the two tissues. Among them, genes involved in immune system/cytokine signalling *(IRF4, CCL19, LTB, JAK3, INPP5D, FCER2, MMP1)* and B cell activation and differentiation (*CD22, BLNK, CARD11*) were over expressed, suggesting an inflammatory signature in the granuloma. Seven granuloma lesion samples clustered with NL tissue samples pointing towards heterogeneity in the granuloma transcriptional profiles (Fig. 2b).

Altogether, our data show a distinct segregation of the granuloma lesion when compared to the NL lung tissue with respect to an inflammatory profile, as previously proposed^11^. Our findings also indicated a diverse range of molecular diversity within the granuloma samples, prompting our decision to delve deeper into the heterogeneity at a transcriptional level.

### Compartments within the granuloma reveal distinct gene expression profiles with an enriched inflammatory response across the lesion

To further explore the granuloma heterogeneity and investigate the contribution of each granuloma compartment to the TB lesion architecture, we first performed an enrichment analysis derived from single sample Gene Set Enrichment analysis (ssGSEA) using the top 40 DEGs discriminating the granuloma from the NL lung tissue. The expression of these genes in the different tissue compartments revealed a more pronounced enrichment score of these DEGs in central and internal lesion samples, suggesting that these two compartments might be the main contributors for the overall granuloma transcriptional signature (Fig. 3a).

**Figure 3.**
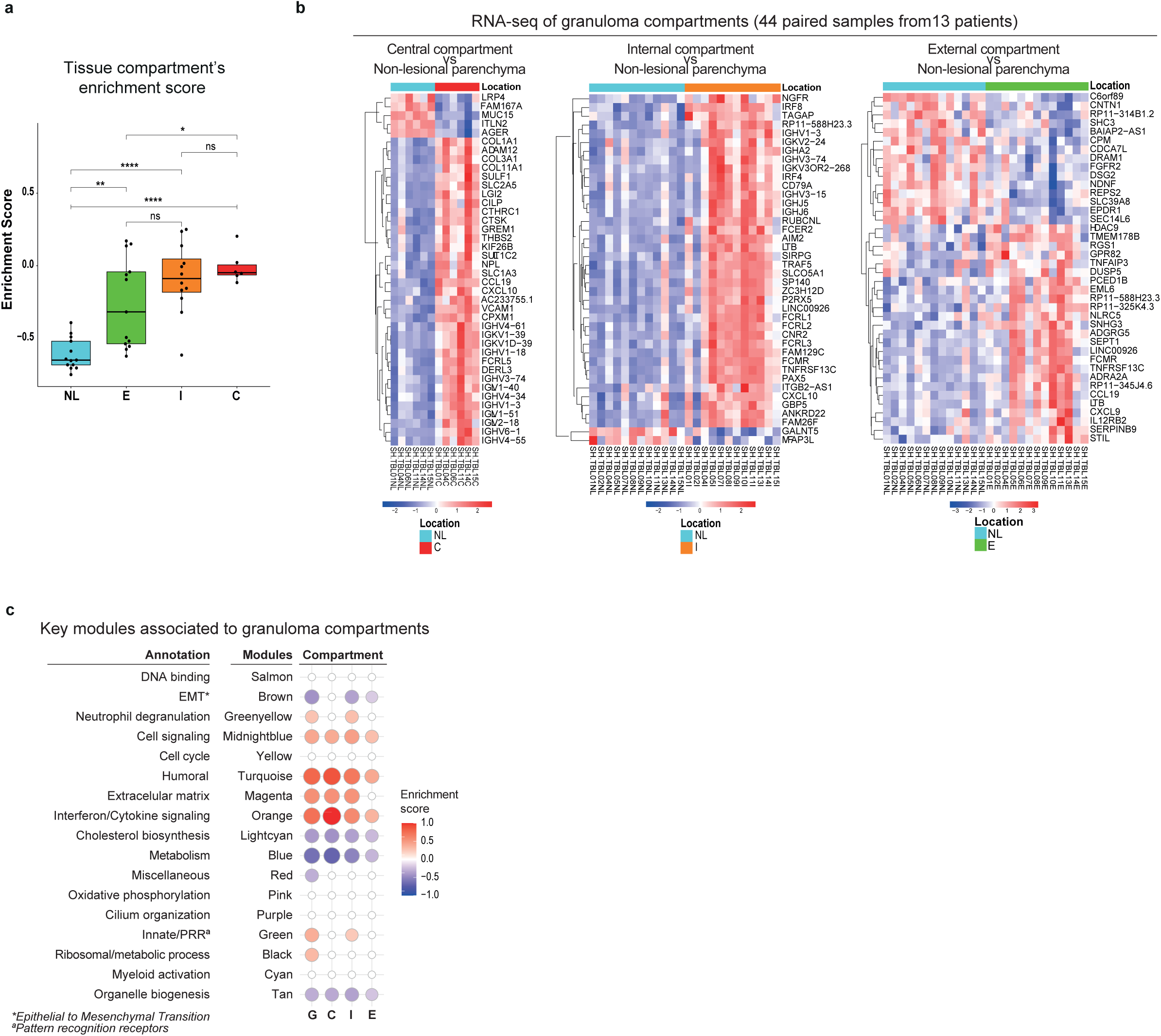
The human lung granuloma compartments have different gene expression profiles and are enriched for immune inflammatory response pathways. **Panel a** show the enrichment score derived from single sample analysis GSEA using the top 40 genes discriminating granuloma from NL lung tissue. Heatmaps showing differences in the top 40 ranked genes from DESeq2 with adjusted p<0.05 by separately comparing the central (C), internal (I) and external (E) compartments with the NL lung tissue gene expression derived **(Panel b).** The intensity of each colour denotes the standardized ratio between each value and the average expression of each gene across all samples. Red pixels correspond to an increased abundance of mRNA in the indicated sample, whereas blue pixels indicate decreased mRNA levels. **Panel c** pictures modular transcriptional of the seventeen modules of co-expressed genes derived from WGCNA for our granuloma dataset separated by compartment. Fold enrichment scores derived using QuSAGE are depicted, with red and blue indicating modules over or under expressed compared to the control. Only modules with fold enrichment (FDR) < 0.1 were considered significant.

Next, we compared the expression profiles derived from each granuloma compartment with the NL tissue. The list of DEGs (DESeq2 with adjusted *p*≤0.05) for the C, I and E *vs* NL tissue comparisons respectively constituted 3,228 (1,539 genes were over-expressed, whereas 1,689 were under-expressed); 5275 (2,676 over-expressed and 2,599 under-expressed); and 1,045 genes (552 over-expressed and 493 under-expressed) (Supplementary Data Set 1 and Extended data Fig. 1b). For central and internal compartments, the hierarchical clustering of the 40 most significantly DEGs showed an evident separation when compared each compartment against the NL lung tissue (Fig. 3b). Though less noticeable, the external compartment was still distinguishable from the NL tissue. Therefore, the magnitude of differential expression relative to NL decreased gradually towards the edge of the granuloma structures (Fig. 3b).

Among the highly variable genes in central lesion, we found genes involved in the immune system/cytokine signalling *(CCL19, CXCL10)* to be upregulated in comparison to the NL tissue (Fig. 3b). On the other hand, we found extracellular matrix organization-related genes to be downregulated (*LRP4, MUC15*), while others were upregulated (*ADAM12, CTSK*). Moreover, collagen-encoding genes (*COL1A1, COL3A1, COL11A1*) were upregulated in the central compartment, which could reflect collagen turnover or fibrosis; as well as of genes associated to immunoglobulin heavy and light chains (*IGHV4−61, IGKV1−39, IGKV1D−39, IGHV1−18, IGHV3−74, IGLV1−40, IGHV4−34, IGHV1−3, IGLV1−51, IGLV2−18, IGHV6−1, IGHV4−55*), related to humoral immunity (Fig. 3b). Furthermore, genes involved in complement fixing (*C1QA, C1QB*, *C1QC*) were significantly upregulated, although not among the top 40 DEGs. For the internal compartment, genes involved in the immune system/cytokine signalling *(LTB, FCMR, AIM2, CXCL10, IRF8, IRF4)* were upregulated compared to NL (Fig. 3b). Furthermore, immune system/cytokine signalling genes *(LTB, CCL19, CXCL9, TNFAIP3, TNFRSF13C, FCMR, AIM2, CXCL10)* were over-expressed in the external compartment relative to NL (Fig. 3b), evidencing an inflammatory signature throughout the granuloma lesion.

We then applied weighted gene co-expression analysis (WGCNA) to perform a modular analysis of co-expressed genes in the granuloma and in the three compartments separately, comparing all samples to NL control tissues. We identified 17 modules from co-expression networks related to the human TB whole granuloma (Fig. 3c, Supplementary Data Set 2). The identified granuloma modular signature showed that neutrophil degranulation, cell signalling, humoral immunity, extracellular matrix, interferon/cytokine signalling, and innate/pathogen recognition receptors (PRR) modules were overabundant. These observations were consistent throughout the compartments, except for the neutrophil degranulation and innate/PRR modules, which were apparent in the total granuloma and the internal lesion only, but not in the central or external lesions (Fig. 3c). Conversely, the Epithelial to Mesenchymal Transition (EMT), cholesterol biosynthesis and metabolism modules were found to be under-abundant in the whole granuloma and external and internal, but not in the central compartment. In addition to cholesterol and metabolism, the organelle biogenesis module was underrepresented, across all compartments, suggesting that some pathways present in the healthy lung are diminished in granuloma tissue (Fig. 3c).

In summary, our results showed a significant enrichment of modules related to inflammation, including pathways of innate immunity in the TB granuloma and the internal compartment, and of humoral immunity across all compartments. Meanwhile a decrease in modules related to extracellular matrix organisation and cholesterol biosynthesis and metabolism was observed in granuloma tissues.

### Patients’ clinical status is associated with differential modular transcriptomic profiles in TB lesions

The heterogeneity in the host immune response to infection, considering the involvement and contribution of physically distinct compartments, together with the bacteria and the inflammatory environment, defines granuloma fate and disease manifestation^16,18^. Hence, we next aimed to associate the modular signature changes in the granulomata (considering the three compartments together) from TB patients with surrogates of treatment response and disease severity. As surrogates of response to treatment we used: achieving stable sputum culture conversion (SCC) before or later than 2 months after the start of anti-TB treatment (fast or slow converters); drug sensitivity of the infecting *Mtb* isolate; whether the individual is a TB relapse or new TB case; and the number of pulmonary lesions present (Supplementary Data Set 2). As surrogates of disease severity, we used presence of symptoms assessed by the St. George’s Respiratory Questionnaire (SGRQ) symptoms score, stratifying individuals according to SGRQ score > 20 or < 20. We quantitatively tested the association of each clinical parameter with each of the significant module’s eigengene (ME) expression patterns (Wilcoxon *p* ≤ 0.05).

Regarding the SCC, the modular signature of the granuloma revealed a significant association of DNA binding and interferon/cytokine signalling modules with SCC, with the enrichment of these modules being significantly higher in those individuals converting the sputum culture later (FDR < 0.1; Figs. 4a and b). No significant modular expression was found to be associated with *Mtb* drug sensitivity of the individuals, relapsed or new cases, or number of lesions (data not shown).

**Figure 4.**
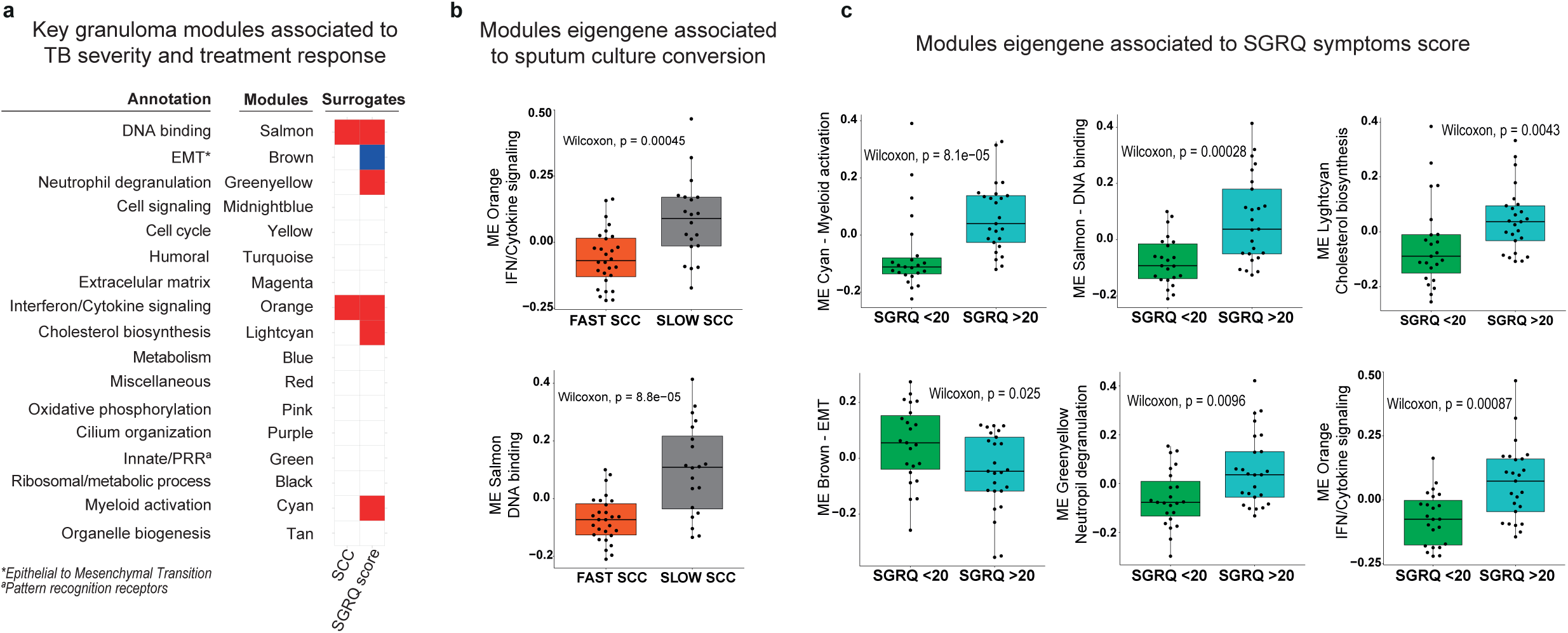
Granuloma modular transcriptional signature correlates with TB clinical and microbiological characteristics revealing differential responses between patient’s group. Modular analysis of granuloma RNA-seq samples from 13 patients. Patients clinically defined accordingly to sputum culture conversion (SCC) and TB disease impact on lung function, measured using the Saint George’s Respiratory Questionnaire (SGRQ) symptom score, as surrogates of TB severity and treatment response. Heatmap represent the key granuloma modules significantly associated to individual’s’ clinical surrogates of TB severity and treatment response **(Panel a).** Fold enrichments were calculated for each WGCNA module using hypergeometric distribution to assess whether the number of genes associated with each clinical status is larger than expected. Fold enrichment scores derived using QuSAGE are depicted, with red and blue indicating modules over or under expressed compared to the control. The colour intensity represents degree of perturbation. Modules with fold enrichment scored FDR p-value < 0.1 are considered significant. **Panel b** and **c** show TB individuals’ stratification according to SCC (fast or slow converters) and SGRQ symptom score (low impact if SGRQ < 20 or high impact if SGRQ > 20), respectively, and the significant association using their corresponding derived WGCNA significant eigengene modules (ME) (p<0.05).

When considering severity of TB disease, we found that DNA binding, neutrophil degranulation, interferon/cytokine signalling, cholesterol biosynthesis and myeloid activation modules were significantly overabundant and associated with higher SGRQ symptoms score (FDR < 0.1; Fig. 4a and c), pointing to higher inflammation status with more severe disease manifestation. In contrast, the EMT module was significantly underabundant in these individuals’ granulomas (Fig. 4c).

Our data from the granulomas of this patient cohort suggest that those patients exhibiting slow SCC, which reflects a poorer response to treatment and slower clearance of *Mtb*, were associated with greater lung inflammatory responses, mainly driven through the interferon/cytokine signalling axis. Additionally, the granuloma signature in patients with the most severe TB cases was also associated with an increased inflammatory response.

## DISCUSSION

The immune response to *Mtb* constitutes a complex and dynamic interaction between the host immune system, the bacteria and lung microenvironment. Throughout infection, the inflammatory response leads to granuloma formation primarily for bacterial containment, while causing extensive tissue remodelling and destruction, which contributes to the clinical spectrum of TB^19^. In our study, we transcriptionally characterised the host response in human pulmonary TB lesions from patients undergoing therapeutic surgery. Fresh human TB lesion specimens obtained from these lung resections (without formalin fixation or paraffin embedding) were transcriptionally profiled using RNA-Seq. Our study provides various advances over previous approaches^15,20,21^. Firstly, we have used a more robust data set with increased patient numbers, which included 44 TB granuloma samples from 13 DS- and MDR-TB patients from the SH-TBL cohort. Secondly, we confirmed a distinct demarcation between granuloma and adjacent non-lesional tissue from the same patient lung. Thirdly, and to our knowledge for the first time, we have identified a transcriptional modular signature within granulomata and linked our findings to clinical/microbiological parameters used as surrogates of TB severity and response to treatment. In patients with more severe disease, our results showed an increased eigengene expression of pro-inflammatory-related modules and a decreased eigengene expression of tissue organization modules. Strikingly, those patients with a delayed response to treatment showed an increased DNA binding and interferon/cytokine modular granuloma response, which has major implications for modifying treatment and subsequent clinical management of disease.

Granuloma heterogeneity in TB is a well-accepted concept and has been reported in non-human primate models of infection and human lesions^16,22^. Besides the heterogeneity among samples, we were able to identify a clear pattern across all TB patients compared to their own NL control lung tissue. Among the top 40 DEGs, we found genes predominantly encoding proteins related to the inflammatory processes that orchestrate the antimycobacterial response such as *CCL19, LTB, FCMR, MMP1* and *IRF4*^23–27^. *CCL19* gene expression was found to be increased in mouse lungs post-*Mtb*-infection to induce lymphoid structures^23^. Regarding human studies, previously published blood microarray-derived signatures found up-regulated levels of *LTB*^28^ when comparing baseline to end-of-treatment samples from TB patients. Moreover, MMP-1 was found to be increased in the respiratory secretions from TB patients and to drive extracellular matrix remodelling in a TB murine model^26^. Recently, *MMP1* was also found to be differentially expressed in TB lymph node biopsies compared to control samples in a study by Reichmann et al^29^. Furthermore, *FCMR* has been considered a target for host-directed therapies^25^, while the transcription factor *IRF4* was found to be required for the generation of Th1 and Th17 subsets of helper T cells and follicular helper T-like cellular responses^27^. In samples from our patient cohort, we found those genes to be over-expressed in the granuloma lesion of patients undergoing surgery for unresolved TB. Additionally, the overexpression of immunoglobulin genes in the granuloma suggests their involvement in complement fixation processes, since *C1QA, C1QB* and *C1QC* transcripts were also found to be upregulated in our granuloma samples. Previously published blood signatures found up-regulated levels of *C1QC*^30,31^, when comparing baseline to end-of-treatment samples. Moreover, the expression of this gene has been proposed as a disease severity biomarker^32^ and linked to poor treatment response^33^. Collectively, these findings suggest that information on the host response within the granulomata from lung resections from critically ill patients, may inform the clinical management of disease.

We explored the granuloma heterogeneity by analysing DEGs between different compartments and applied WGCNA modular analysis to the whole granuloma and separated compartments. In line with Dheda’s and Marakalala’s studies^11,16^, we showed that both the centre and internal compartments of the TB granuloma lesion present a more perturbed transcript abundance compared to the external lesion compartment, potentially pointing to the trajectory of the host response to *Mtb*. Our results confirm the involvement of pro-inflammatory-related genes and modules in the human TB granuloma and compartments. The spatial differences within the granuloma in terms of gene expression are supported by other studies^11,15^. Dheda et al. described the pathways involved in different parts of cavitary lesions from 14 failed MDR-TB subjects that underwent surgery, pointing to the cavity wall as the main source of pro-inflammatory activity^11^. In line with our findings, they showed that pro-inflammatory pathways were especially over-represented in the cavity wall, including nitric oxide production, reactive oxygen species, IL-1, IL-6, IFN-γ and NF-κβ transcriptional signatures. Furthermore, Subbian et al. demonstrated using four granuloma samples, the involvement of immune cell signalling and activation, interferon response and tissue remodelling processes in the complex TB granuloma microenvironment^15^. The modular granuloma signature that we describe herein, provides comparable and additional data, albeit in a larger and independent patient cohort.

We identified 17 modules from co-expressed networks and mapped a TB granuloma modular signature, consisting of increased neutrophil degranulation, cell signalling, humoral immunity, extracellular matrix, interferon/cytokine signalling and innate/PRR. In our cohort, patients presented advanced TB disease with cavitary TB. We found the neutrophil module to be increased in whole granuloma but not in the central or external lesions, possibly explained by more necrosis in this region, coupled with relatively low RNA abundance in neutrophils. Moreover, the *MMP1* gene was overexpressed in the granuloma as compared to NL tissue, which might suggest the involvement of neutrophils through the activity of matrix metalloproteinases. In humans, neutrophil accumulation in the lungs of active TB patients and has been correlated with increased lung pathology and consequent disease progression^34,35^. The role of neutrophils in TB disease progression and pathology has been well documented in experimental mouse models^36–39^. Additionally, the extracellular matrix, interferon/cytokine signalling and innate/PRR modules have been reported to be upregulated in blood from TB patients^40,41^, demonstrating that whatever is happening at the site of infection can also be translated to the periphery. Interestingly, we found that the humoral module was increased in whole granuloma samples, corroborating with the expression of immunoglobulin heavy and light chains transcripts in both central and internal compartments. Our findings are in keeping with a report from Krause et al., demonstrating the presence of different B cell subsets and high levels of *Mtb*-reactive antibodies in human lung tissue^42^.

Conversely, the EMT, cholesterol biosynthesis and metabolism modules were decreased in granuloma relative to NL parenchyma. The EMT is linked to wound healing but also to fibrogenesis and scarring^43^. The downregulation of this module in granuloma, along with its decreased enrichment in more severe TB cases compared to those with milder symptoms, might suggest a disruption in critical processes needed for tissue repair. We also observed increased cholesterol synthesis in individuals experiencing more severe disease in terms of presenting more pronounced symptomatology. Kim et al. proposed dysregulation of host lipid metabolism caused by *Mtb*, tracing the progression of TB granulomas to caseation, cavitation, and eventual disease transmission^20^. The authors suggested that bacterial components could trigger the host’s innate immune system, potentially augmenting the synthesis or storage of host lipids. Consequently, in line with these findings^20^ the upsurge in cholesterol synthesis which we observe herein might mirror the impact of *Mtb* on the host lipid metabolism.

Indeed, a major novelty of our work is the use of unbiased modular analysis to link the transcriptional signature generated from TB granuloma lesions with patients’ clinical surrogates of TB severity and the time taken to clear *Mtb* in sputum, and thus response to treatment. Specifically, our results show an important inflammation component in granulomas from patients presenting with greater severity of disease and slower response to treatment. Inflammation has been described as key for tissue damage and linked to a blood transcriptional signature in individuals suffering from active TB disease even months before being diagnosed^44^, and radiographic lung disease extension^7,36,40^, decreasing upon treatment^40,45^. Tabone et al. revealed differential responses in the blood transcriptional signature among various clinical TB subgroups following treatment, observing a reduction in the inflammation and IFN modules alongside B and T cell modular signatures accompanying successful treatment^41^. This observation raises the logical assumption that changes induced by treatment at the blood level might parallel alterations occurring at the infection site. In our study, we found an overabundance of the IFN/cytokine signalling and DNA binding modules associated with severe disease, characterized by worsened symptoms and slower bacterial clearance. Our observations also hint at a potential connection between heightened neutrophil degranulation in severe cases and the damaging mechanisms associated with neutrophil-mediated inflammation^46^, suggesting a plausible role for this process in exacerbating the severity of the condition. As our study samples were collected after the end of treatment, all cases examined here could be considered difficult or inadequate responders to treatment. This inadequate response may result in a sustained pro-inflammatory profile at the granuloma site or be the consequence of it, and our findings may thus help in future management of disease treatment. Both our data and data from other sources collectively offer the opportunity to use these transcript patterns to monitor treatment response.

Our results suggest that the clinical picture mirrors the inflammation happening at the site of infection and confirms what has been previously seen by others indirectly, both in humans and in experimental animal models. Malherbe et al., showed through 18F-FDG-PET-CT lung scans that some patients still have an increased FDG uptake in the lesions when compared to surrounding healthy tissue after six months of treatment^8^, and more recently, the authors have related both a larger burden of disease and a slower rate of reduction in scan metrics with delayed sputum coversion^47^. Our data showed that slow sputum converters present different modular expression profiles in TB granulomas when compared to fast sputum converters. To date, the SCC constitutes the only tool endorsed by the WHO to monitor treatment response^48^ and can be considered a surrogate of disease severity. Therefore, achieving SCC after two months of starting treatment has been associated with TB cavity persistence^14^ and poor prognosis^49,50^. In our cohort, the proportion of DS-TB/MDR-TB cases in the fast SCC converters was higher (5/8) than in slow converters (4/6), as previously described^48^. Importantly, from our findings, the analysis of granulomatous tissue in resected lung samples from patients suggests that the slower SCC converters exhibit the highest expression of inflammatory genes in the lesion after several months, supporting the potential use of SCC at the two-month mark after treatment initiation as a prognostic indicator. On the other hand, time to sputum clearance could enhance treatment monitoring and refine clinical management and might assist in identifying patients who should be prioritized for further therapeutic interventions. Interesting, measures of microbiological treatment success and clinical severity of disease have also been associated with *Mtb* transcriptional profiles in patient sputa^51^, suggesting that the lesion immunopathology described here also impact *Mtb* lung phenotypes.

Our study has some limitations. These TB individuals presented advanced TB disease with cavitary TB, and underwent lung resection surgery, so the results may not be generalisable to the whole spectrum of TB disease. In countries with a high prevalence of MDR-TB, adjunctive surgical resection is a common therapeutic tool which, despite being a major invasive procedure, reduces the transmission burden of MDR-TB and results in favourable outcomes for the patients^52^. However, this approach is uncommon in most countries, thus our results help to understand TB host response but may have a direct impact on TB treatment at short term only in high burden countries where resection is practised. Furthermore, given that individuals with TB may have several lesions at varying stages, which can evolve and recede (as shown in experimental animal studies^4,7,53,54^), expanding the sample size to include several lesions from the same individuals would be beneficial. However, achieving this is practically unfeasible without conducting a complete pulmonectomy or lung section resection. Consequently, working with samples collected *post-mortem* could offer a viable solution, offering substantial insights and information in this regard, although this is limited by the number of TB patients from whom *post-mortem* samples would be available and the quality of the samples collected.

In conclusion, we have defined a robust signature for human advanced TB granuloma lesions, despite the inter granulomata heterogeneity. Moreover, this is the first study showing different modular transcriptomic signature patterns, integrating and co-analysing our findings with TB patients’ clinical/microbiological measurements, including severity and response to treatment. Our study provides a considerable dataset on TB granulomas gene expression which will undoubtedly be of broad utility, interest and significance to the scientific community, contributing to an increase in knowledge on TB immunopathology. A better understanding of disease processes and host protective immune responses may help in the clinical management of TB and development of treatment strategies. Most importantly, our findings provide evidence of the clinical picture with a relationship between clinical parameters, treatment response and immune signatures at the infection site.

## METHODS

### Ethics

This study is part of the SH-TBL project (ClinicalTrials.gov Identifier: NCT02715271). The protocol, research methodology and all associated documents (informed consent sheet, informed consent form) were reviewed and approved by both ethics’ committees at the National Center of Tuberculosis and Lung Diseases (NCTLD) (IRB00007705 NCTLD Georgia #1, IORG0006411) and the Germans Trias i Pujol University Hospital (EC: PI-16-171). Written informed consent was obtained from all study participants before enrolment.

### Study design and patient cohort

Study participants constituted the SH-TBL cohort, a cross-sectional study recruiting 40 adult patients was conducted in the NCTLD in Tbilisi (Georgia), from May 2016 to May 2018, after receiving therapeutic surgery indication for their pulmonary TB. All volunteers received standard anti-TB treatment (ATT) regimen according to Georgia national guidelines and achieved sputum culture conversion. Surgery was required due to persistent radiological signs of cavitary lesions on chest X-Ray and computed tomography scan, according to the Georgian National Guideline “Surgical Treatment of Patients with Pulmonary Tuberculosis” based on official guidelines of surgery recommendations for pulmonary TB. Thoracic surgery decisions were made by the NCTLD Resistant Tuberculosis Treatment Committee, composed of two surgeons and 18 pulmonary TB specialists.

### Data and sample collection

Anonymised data regarding the socio-epidemiological factors, clinical aspects, and information referring to the current TB episode for the SH-TBL cohort were collected using an electronic case report form. TB individuals were classified taking into consideration clinical surrogates of disease severity. According to their sputum conversion date, individuals were categorized as fast (SCC < 2 months) or slow converters (SCC > 2 months). Drug sensitivity of the *Mtb-*infecting isolate was recorded, classifying TB cases as DS or MDR-TB. Presence of symptoms was assessed with the SGRQ symptom score, and a cut off defined by the median SGRQ symptoms value (SGRQ cut off = 20) was used to stratify the individuals: those with an SGRQ score > 20 were considered to have more severe TB disease than those with SGRQ score < 20. Relapsed cases and number of lesions were also considered (Supplementary Data Set 2 and supplementary Tables 1 and 2). During surgery, TB granuloma lesions of 3.2 cm in diameter (median; 2.2-5.5 cm) were removed according to surgical criteria. In total, 48 SH-TBL biological samples were collected for the study. Biopsies from three different sections of TB lesion were collected, namely Central Lesion (C), Internal Wall (I), External Wall (E). In addition, surrounding non-lesional (NL) lung parenchyma tissue, unaffected, by eye and by palpation, was collected from the same patient (Fig. 2a). Biopsy fragments (∼0.5 cm^3^) of tissue samples from 14 patients were collected for RNA-Seq. Fresh tissue samples were incubated in RNA*later* solution (Qiagen) at 4°C overnight, before storage at -80°C. Samples were processed in BioSafety Level 3 (BSL-3) laboratory.

### Total RNA extraction

For an optimal RNA recovery, TB lesion biopsy samples were divided into 0.16-0.21g single pieces and placed into new tubes. Samples were reduced to powder by mechanical cryofracturing using a BioPulverizer device (Biospec Products) after being cooled in liquid nitrogen. The powdered tissue was then transferred to 2 mL Lysing Matrix D tubes together with lysis solution for homogenization by FastPrep® instrument (MP Biomedicals). RNA was purified using the mirVana miRNA Isolation Kit (Thermo Fisher Scientific), followed by genomic DNA digestion using the DNA-*free* DNA Removal Kit (Thermo Fisher Scientific) according to manufacturer’s instructions. Quantitative and qualitative RNA integrity number equivalent (RINe) values were obtained by Agilent Bioanalyzer 2100 (Agilent Technologies). In general, a standard RINe score for good quality RNA is set at seven. Considering the source and status of the human TB lesion samples, a minimum RINe cut-off of four was established, accepting that low RNA integrity is a potential source of bias in RNA-Sequencing (RNA-Seq) experiments.

### RNA-Sequencing library preparation, sequencing, and gene alignment

Purified RNA was diluted to 25 ng/µl per aliquot and then shipped on dry ice to Macrogen (Seoul, South Korea), where the RNA-sequencing (RNA-Seq) was performed. Libraries were constructed using the TruSeq Stranded Total RNA LT Sample Prep Kit (Human Mouse Rat) (Illumina) following the TruSeq Stranded Total RNA Sample Prep guide (Part #15031048 Rev. E), including prior removal of ribosomal RNA using the RNA Ribo-Zero rRNA Removal Kit (Human/Mouse/Rat) (Illumina). RNA-Seq was performed on an Illumina platform HiSeq 4000 (Illumina), at 50 million reads per sample, 100bp stranded paired-end reads. Pre-processing of raw data included quality control through FastQC (v.011.7) and MultiQC (v.1.9)^55^. Before further steps in read pre-processing, Illumina adapters were trimmed off with Trimmomatic (v.0.39)^56^. The human genome sequence GRCh38.89 and human gene annotations were downloaded from the ENSEMBL web repository. Files from each sample were aligned to the human reference genome using the Spliced Transcripts Alignment to a Reference (STAR) package (v.2.7.5b)^57^, with the built-in gene counts quantification mode for stranded RNA-Seq data. BAM files were generated, and the SAMtools package applied to calculate the percentage of successful read alignment against the reference human genome (v.1.10)^57^

### RNA-seq data analysis

The overall pipeline for data handling, plotting and statistical analysis was conducted in R (v.3.6.2). After STAR mapping, a gene count data table was obtained including C, I, E, and NL samples. Genes with a lower than 50 counts among all the samples were discarded to avoid confounding the differential gene expression analysis, as they had low expression to be reliably quantified. Paired statistical analyses were done globally and separately for each compartment. The set for the RNA-Seq experiments comprised 44 paired samples from 13 patients. Samples from patient SH-TBL03 weren’t taken into consideration for the RNA-Seq analysis as the NL tissue sample control was missing. The differential expression analysis from tissue count tables was conducted using the DESeq2 Bioconductor package (v.1.28.1)^58^. Genes were considered as significant DEGs when the Benjamini-Hochberg adjusted *p*-value was equal to or less than 0.05 (*p* ≤ 0.05). The R package heatmap (v.1.0.12) was used to generate heatmaps and dendrograms for the genes and samples by hierarchical clustering after DESeq2 depth normalization. Heatmaps describe the Euclidean distances between samples.

### Enrichment score for the different tissue compartments

The expression across compartments of upregulated selected genes differentiating granuloma from non-lesional tissue was performed using single sample Gene Set Enrichment analysis (ssGSEA). ssGSEA is a variation of the GSEA algorithm that instead of calculating enrichment scores for groups of samples and sets of genes, it provides a score for each sample and gene set pair^59^.

### Weighted gene co-expression network analysis and functional annotation

Weighted gene co-expression network analysis (WGCNA) was performed to identify modules using the R package WGCNA. The TB granuloma modules were constructed using the 10,000 most variable genes across all TB samples collected (log2 RNA-seq expression values). To satisfy the scale-free topology criteria and the recommendations for WGCNA use, we chose an optimal soft-threshold (β = 22.5) to obtain an adjacency matrix from a signed weighted correlation matrix containing pairwise Pearson correlations, generating the corresponding topological overlap measure (TOM). To detect the modules, we applied a dynamic hybrid tree-cut algorithm to detect the computed modules of co-expressed genes (minimum module size of 20, and deep split = 1). Finally, 21 modules were obtained. An additional “grey” module was identified in TB granuloma modules, consisting of genes that were not co-expressed with any other genes. The grey module was discarded from further analysis. Moreover, only modules with more than 40 genes were annotated. We computed their intramodular connectivity and selected the top five most interconnected genes^60^. Significantly enriched Gene Ontology and canonical pathways from the MSigDB website^61^ were computed using clusterProfiler R package^62^. Modules were annotated based on representative biological processes from pathways and processes from all three reference databases. Fold enrichment for the WGCNA modules was calculated using the quantitative set analysis for gene expression with the Bioconductor package QuSAGE^63^. To identify the modules of genes over- or under-abundant in TB granuloma, compared to the respective non-lesional lung tissue using log2 expression values using the three compartments. Only modules with enrichment scores with FDR *p*-value < 0.1 were considered significant.

### Association between modules and clinical characteristics

The following parameters were used as surrogates of TB severity: presence of symptoms assessed by the SGRQ symptoms score; achieving the SCC before or later than 2 months after the start of anti-TB treatment; DS vs MDR-TB case; being a relapse or new TB case; and number of lesions present. We computed the eigengene for each module, defined as the first principal component of the module representing the overall expression level of the module. The relationship of the transcriptomic modules with clinical surrogates of TB severity (SCC and SGRQ symptoms score) was tested using Wilcoxon-rank sum test. Nominal *p*-values were adjusted using the Benjamini-Hochberg approach^64^.

## ACKNOWLEDGEMENTS

The authors would like to thank the patients who agreed to participate in the study and the staff from the NCTLD who helped with the SH-TBL project. The authors thank the support from Comparative Medicine and Bioimage Centre of Catalonia (CMCiB), namely at BSL 3 facility, Eric Garcia for technical support in acquiring the data. The authors, thank William J. Branchett, from The Francis Crick Institute, London, for his input on the manuscript and providing suggestions/changes for improvement.

## AUTHOR CONTRIBUTIONS

Conception: CV and SV. Design of the work: CV and SV, with substantial contributions of MK and AOG. KLF, AD, JJL, JS, LA, DHC, Ad R, JCR, AG, LMW, PJC, SV, SG, KN, NS and ZA worked on the acquisition and analysis of data for the work, and all authors made substantial contributions to its interpretation. RNA-Sequencing library preparation, sequencing, gene alignment and initial bioinformatics analysis was performed by AD and DHC, supervised by MK; and paired statistical analysis, single sample Gene Set Enrichment analysis, WGCNA analysis and the association between modules and clinical characteristics were performed by KLF and JJL, supervised by AOG. KLF and CV drafted the work; and CA, FMT, AS, AGC, SV, SW, MK and AOG reviewed it critically for important intellectual content. All authors revised and gave their final approval of the version to be published and agreed to be accountable for all aspects of the work in ensuring that questions related to the accuracy or integrity of any part of the work are appropriately investigated and resolved.

## DATA AVAILABILITY

Patient’s data were transferred to a data frame (SH-TBL Cohort metadata) and are freely accessible (doi: 10.17632/knhvdbjv3r.1). All metadata and sequencing data from this manuscript have been deposited in the National Center for Biotechnology Information Gene Expression Omnibus (GEO) database and will be released after manuscript acceptance in a peer-reviewed scientific journal.

## Extended Data Figure

**Extended data Figure 1.**
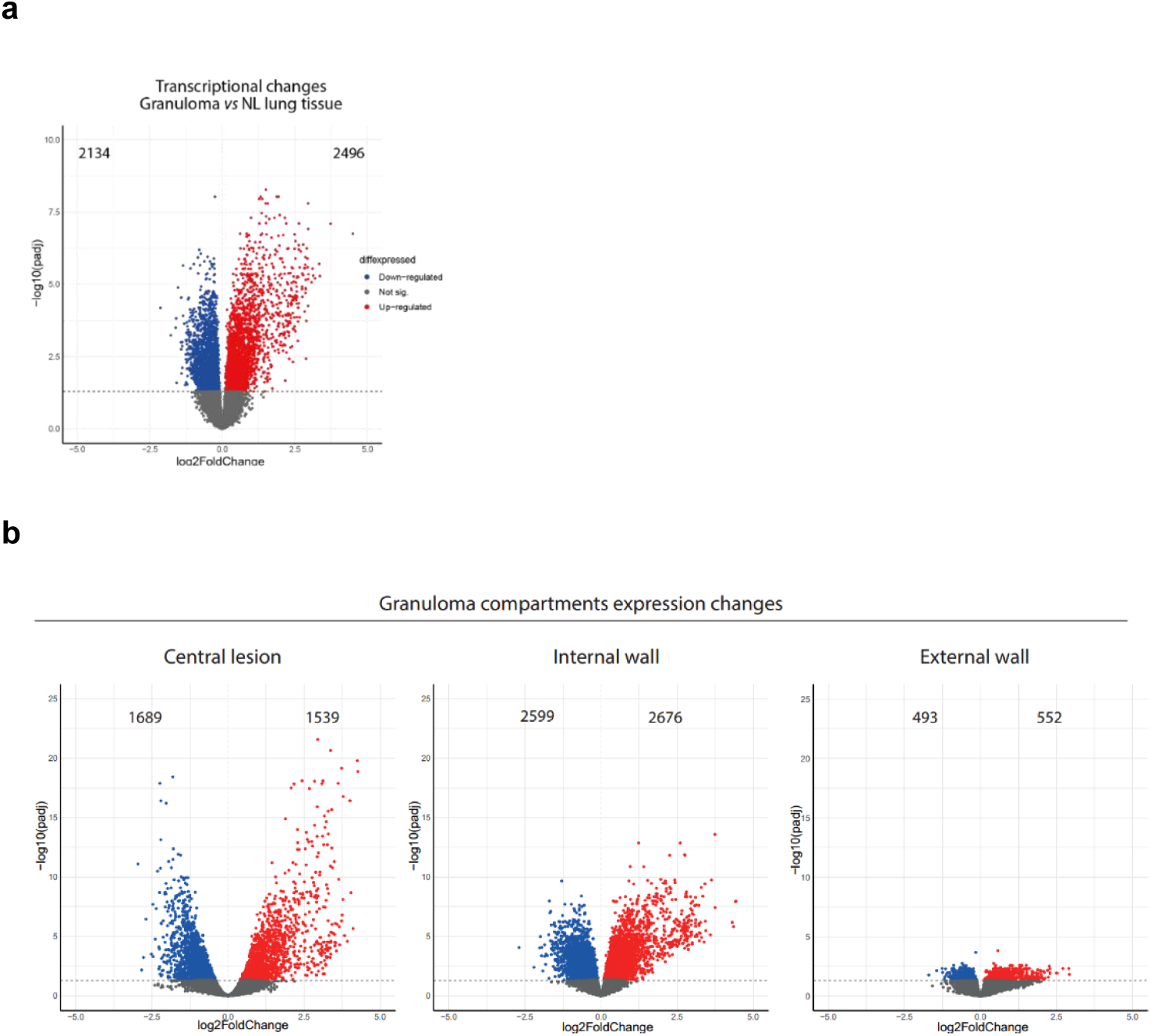
Differential expression analysis of human TB lung granuloma and separated compartments relative to non-lesional lung tissue. **Panel a)** Volcano plot depicts differentially expressed genes (DEGs) between granuloma and non-lesional (NL) lung tissue. **Panel b)** Volcano plot depicts DEGs between Central, Internal and External compartments and the non-lesional (NL) lung tissue. Genes with the adjusted *p*-value ≤0.05 were considered significantly down-regulated when the log2 fold change was < 0.1 (blue) and up-regulated when log2fold change > 0.1 (red).

## Supplementary Information

**Supplementary Table 1.** Demographic and clinical and TB-related patients’ characteristics at time of sur1g4ery.

| Variable | Category | Overall | Male | Female | *p*-value |
| --- | --- | --- | --- | --- | --- |
| *N* |  | *N*= 14 | *n*= 7 | *n*= 7 |  |
| Clinical & epidemiological characteristics |  |  |  |  |  |
| Age [mean (SD)] |  | 34.36 (12.02) | 37 (15.56) | 31 (7.39) | 0.4387 [a] |
| BMI [median (range)] |  | 23.55 [19.1-32] | 23.4 [21.5-31-7] | 23.7 [19.1-32] | 0.6841 [b] |
| Smoker (%) | Yes | 4 (28.57) | 4 (57.14) | 0 (0) | 0.0699 [c] |
|  | No | 10 (71.43) | 3 (42.86) | 7 (100) |  |
| Alcohol (%) | Yes | 5 (35.71) | 5 (71.43) | 0 (0) | 0.0210* [c] |
|  | No | 9 (64.29) | 2 (28.57) | 7 (100) |  |
| Comorbidities (HCV, HBV, Diabetes) (%) | Yes | 6 (42.86) | 3 (42.86) | 3 (42.86) | 1 [c] |
|  | No | 8 (57.14) | 4 (57.14) | 4 (57.14) |  |
| Characteristics of current TB episode |  |  |  |  |  |
| Patient history (%) | Relapse | 5 (37.71) | 2 (28.57) | 3 (42.86) | 1 [c] |
|  | New patient | 9 (64.29) | 5 (71.43) | 4 (57.14) |  |
| Drug Sensitivity (%) | DS-TB | 7 (50) | 3 (42.86) | 3 (42.86) | 1 [c] |
|  | MDR/XDR-TB | 7 (50) | 4 (57.14) | 4 (57.14) |  |
| Fast (≤2 months) and slow (>2 months) time to sputum culture conversion | Fast converters | 8 (57.14) | 3 (42.86) | 5 (71.43) | 0.5921 [c] |
|  | Slow converters | 6 (42.86) | 4 (57.14) | 2 (28.57) |  |
| Multiple lesions in the CXR | Yes (≥2) | 4 (28.57) | 2 (28.57) | 2 (28.57) | 1 [c] |
|  | No (<2) | 10 (71.43) | 5 (71.43) | 5 (71.43) |  |
| Macroscopic and microbiological characteristics of the TB lesions |  |  |  |  |  |
| Size of resected lesions (mm) [median (range)] |  | 30 [29-42] | 30 [30-35] | 30 [29-42] | 0.4371 [b] |
| Necrosis (%) | Fresh | 13 (92.86) | 6 (85.71) | 7 (100) | 1 [c] |
|  | Not fresh necrosis | 1 (7.14) | 1 (14.29) | 0 (0) |  |
| Granuloma biopsy culture conversion by patient (%) | Positive | 1 (17.14) | 1 (14.29) | 0 (0) | 1 [c] |
|  | Negative | 13 (92.86) | 6 (85.71) | 7 (100) |  |

Statistics: [a] t-test, [b] Mann-Whitney U-test, [c] Fisher's exact test. \*Statistically significant differences found in alcohol use only).

**Supplementary Table 2.** Characteristics of resected lesions from TB patients according to treatment response.

| Variable | Category | Overall | Fast Responders | Slow Responders | *p*-value |
| --- | --- | --- | --- | --- | --- |
| *N* |  | *N* = 14 | *n* = 8 | *n* = 6 |  |
| Size of resected lesion (mm) [median (range)] |  | 30 [29-42] | 30 [29-42] | 32.5 [30-40] | 0.5035 [a] |
| Necrosis (%) | Fresh necrosis | 13 (92.86) | 8 (100) | 5 (83.33) | 0.4286 [b] |
|  | Not fresh necrosis | 1 (7.14) | 0 (0) | 1 (16.67) |  |
| Cavitation (%) | Cavitation | 11 (78.57) | 5 (62.5) | 6 (100) | 0.2088 [b] |
|  | Tuberculoma | 3 (21.43) | 3 (37.5) | 0 (0.00) |  |
| Biopsy culture positivity |  |  |  |  |  |
| Biopsy culture conversion by patient (%) | Positive | 1 (7.14) | 0 (0.00) | 1 (16.67) | 0.4286 [b] |
|  | Negative | 13 (92.86) | 8 (100) | 5 (83.33) |  |

Statistics: [a] Mann-Whitney U-test, [b] Fisher's exact test.
Comparisons refer to patients presenting microbiological conversion at two months or less against those who required more than two months to respond to standard anti-TB treatment.

## REFERENCES

1. World Health Organization (WHO). Global Tuberculosis Report. (2022).

2. World Health Organization (WHO). WHO Report on TB 2020. (2020).

3. World Health Organization. Global Tuberculosis Report. (2021).

4. Gil, O. et al. Granuloma encapsulation is a key factor for containing tuberculosis infection in minipigs. PLoS One 5, e10030 (2010).

5. Lenaerts, A., Barry, C. E. & Dartois, V. Heterogeneity in tuberculosis pathology, microenvironments and therapeutic responses. Immunol Rev 264, 288–307 (2015).

6. Lin, P. L. et al. Sterilization of granulomas is common in active and latent tuberculosis despite within-host variability in bacterial killing. Nat Med 20, 75–79 (2014).

7. Gideon, H. P. et al. Multimodal profiling of lung granulomas in macaques reveals cellular correlates of tuberculosis control. Immunity 55, 827–846 (2022).

8. Malherbe, S. T. et al. Persisting PET-CT lesion activity and M. tuberculosis mRNA after pulmonary tuberculosis cure. Nat Med 22, 1094 (2016).

9. Penn-Nicholson, A. et al. RISK6, a 6-gene transcriptomic signature of TB disease risk, diagnosis and treatment response. Sci Rep 10, 8629 (2020).

10. Urbanowski, M. E., Ordonez, A. A., Ruiz-Bedoya, C. A., Jain, S. K. & Bishai, W. R. Cavitary tuberculosis: the gateway of disease transmission. Lancet Infect Dis 20, e117–e128 (2020).

11. Dheda, K. et al. Spatial network mapping of pulmonary multidrug-resistant tuberculosis cavities using RNA sequencing. Am J Respir Crit Care Med 200, 370– 380 (2019).

12. Kempker, R. R. et al. Additional drug resistance in Mycobacterium tuberculosis isolates from resected cavities among patients with multidrug-resistant or extensively drug-resistant pulmonary tuberculosis. Clinical Infectious Diseases 54, 51–54 (2012).

13. Dartois, V. & E. Barry, C. Clinical pharmacology and lesion penetrating properties of second- and third-line antituberculous agents used in the management of multidrug-resistant (MDR) and extensively-drug resistant (XDR) tuberculosis. Curr Clin Pharmacol 5, 96–114 (2010).

14. Vashakidze, S. et al. Retrospective study of clinical and lesion characteristics of patients undergoing surgical treatment for Pulmonary Tuberculosis in Georgia. Int J Infect Dis 56, 200–207 (2017).

15. Subbian, S. et al. Lesion-specific immune response in granulomas of patients with pulmonary tuberculosis: A pilot study. PLoS One 10, 1–21 (2015).

16. Marakalala, M. J. et al. Inflammatory signaling in human tuberculosis granulomas is spatially organized. Nat Med 22, 531–538 (2016).

17. McCaffrey, E. F. et al. The immunoregulatory landscape of human tuberculosis granulomas. Nature Immunology 23, 318–329 (2022).

18. Lin, P. L. & Flynn, J. L. The End of the Binary Era: Revisiting the Spectrum of Tuberculosis. The Journal of Immunology 201, 2541–2548 (2018).

19. McCaffrey, E. F. et al. The immunoregulatory landscape of human tuberculosis granulomas. Nat Immunol 23, 318–329 (2022).

20. Kim, M. J. et al. Caseation of human tuberculosis granulomas correlates with elevated host lipid metabolism. EMBO Mol Med 2, 258–274 (2010).

21. McCaffrey, E. F. et al. The immunoregulatory landscape of human tuberculosis granulomas. Nature Immunology 23, 318–329 (2022).

22. Cadena, A. M., Fortune, S. M. & Flynn, J. L. Heterogeneity in tuberculosis. Nature Reviews Immunology 17, 691–702 (2017).

23. Kahnert, A. et al. Mycobacterium tuberculosis triggers formation of lymphoid structure in murine lungs. Journal of Infectious Diseases 195, 46–54 (2007).

24. Agyekum, S. et al. Expression of lymphotoxin-beta (LT-beta) in chronic inflammatory conditions. J Pathol 199, 115–121 (2003).

25. Wang, H., Coligan, J. E. & Morse, H. C. Emerging Functions of Natural IgM and Its Fc Receptor FCMR in Immune Homeostasis. Front Immunol 7, (2016).

26. Elkington, P. et al. MMP-1 drives immunopathology in human tuberculosis and transgenic mice. J Clin Invest 121, 1827 (2011).

27. Swanson, R. V. et al. Antigen-specific B cells direct T follicular-like helper cells into lymphoid follicles to mediate Mycobacterium tuberculosis control. Nature Immunology 24, 855–868 (2023).

28. Cliff, J. M. et al. Distinct phases of blood gene expression pattern through tuberculosis treatment reflect modulation of the humoral immune response. J Infect Dis 207, 18–29 (2013).

29. Reichmann, M. T. et al. Integrated transcriptomic analysis of human tuberculosis granulomas and a biomimetic model identifies therapeutic targets. Journal of Clinical Investigation 131, (2021).

30. Bloom, C. I. et al. Detectable Changes in The Blood Transcriptome Are Present after Two Weeks of Antituberculosis Therapy. PLoS One 7, e46191 (2012).

31. Ottenhoff, T. H. M., Dass, R. H., Yang, N., Zhang, M. M. & Wong, H. E. E. Genome-Wide Expression Profiling Identifies Type 1 Interferon Response Pathways in Active Tuberculosis. PLoS One 7, 45839 (2012).

32. Cai, Y. et al. Increased complement C1q level marks active disease in human tuberculosis. PLoS One 9, (2014).

33. Cliff, J. M. et al. Distinct phases of blood gene expression pattern through tuberculosis treatment reflect modulation of the humoral immune response. J Infect Dis 207, 18–29 (2013).

34. Eum, S.-Y. et al. Neutrophils Are the Predominant Infected Phagocytic Cells in the Airways of Patients With Active Pulmonary TB. Chest (2010).

35. Gopal, R. et al. S100A8/A9 proteins mediate neutrophilic inflammation and lung pathology during tuberculosis. Am J Respir Crit Care Med 188, 1137–1146 (2013).

36. Moreira-Teixeira, L. et al. Mouse transcriptome reveals potential signatures of protection and pathogenesis in human tuberculosis. Nature Immunology 2020 21:4 21, 464–476 (2020).

37. Fonseca, K. L. et al. Deficiency in the glycosyltransferase Gcnt1 increases susceptibility to tuberculosis through a mechanism involving neutrophils. Mucosal Immunology 13, 836–848 (2020).

38. Nandi, B. & Behar, S. M. Regulation of neutrophils by interferon-γ limits lung inflammation during tuberculosis infection. J Exp Med 208, 2251–2262 (2011).

39. Dorhoi, A. et al. The adaptor molecule CARD9 is essential for tuberculosis control. J Exp Med 207, 777–792 (2010).

40. Berry, M. P. R. et al. An interferon-inducible neutrophil-driven blood transcriptional signature in human tuberculosis. Nature 466, 973–977 (2010).

41. Tabone, O. et al. Blood transcriptomics reveal the evolution and resolution of the immune response in tuberculosis. Journal of Experimental Medicine 218, (2021).

42. Krause, R., et al. B cell heterogeneity in human tuberculosis highlights compartment-specific phenotype and putative functional roles. PREPRINT (Version 1) available at Research Square (2023).

43. Stone, R. C. et al. Epithelial-Mesenchymal Transition in Tissue Repair and Fibrosis. Cell Tissue Res 365, 495 (2016).

44. Scriba, T. J. et al. Sequential inflammatory processes define human progression from M. tuberculosis infection to tuberculosis disease. PLoS Pathog 13, 1–24 (2017).

45. Thompson, E. G. et al. Host blood RNA signatures predict the outcome of tuberculosis treatment. Tuberculosis 107, 48–58 (2017).

46. Tiwari, D. & Martineau, A. R. Inflammation-mediated tissue damage in pulmonary tuberculosis and host-directed therapeutic strategies. Semin Immunol 65, 101672 (2023).

47. Malherbe, S. T. et al. Quantitative 18F-FDG PET-CT scan characteristics correlate with tuberculosis treatment response. EJNMMI Res 10, 1–15 (2020).

48. Kurbatova, E. V. et al. Sputum culture conversion as a prognostic marker for end-of-treatment outcome in patients with multidrug-resistant tuberculosis: a secondary analysis of data from two observational cohort studies. Lancet Respir Med 3, 201–209 (2015).

49. Wallis, R. S. et al. Biomarkers for tuberculosis disease activity, cure, and relapse. Lancet Infect Dis 9, 162–172 (2009).

50. Assemie, M. A., Alene, M., Petrucka, P., Leshargie, C. T. & Ketema, D. B. Time to sputum culture conversion and its associated factors among multidrug-resistant tuberculosis patients in Eastern Africa: A systematic review and meta-analysis. Int J Infect Dis 98, 230–236 (2020).

51. Honeyborne, I. et al. Profiling persistent tubercule bacilli from patient sputa during therapy predicts early drug efficacy. BMC Med 14, (2016).

52. Kempker, R. R., Vashakidze, S., Solomonia, N., Dzidzikashvili, N. & Blumberg, H. M. Surgical treatment of drug-resistant tuberculosis. Lancet Infect Dis 12, 157– 166 (2012).

53. Hunter, L., Hingley-Wilson, S., Stewart, G. R., Sharpe, S. A. & Salguero, F. J. Dynamics of Macrophage, T and B Cell Infiltration Within Pulmonary Granulomas Induced by Mycobacterium tuberculosis in Two Non-Human Primate Models of Aerosol Infection. Front Immunol 12, 776913 (2022).

54. Canetti, G. The Tubercle Bacillus in the Pulmonary Lesion of Man: Histobacteriology and Its Bearing on the Therapy of Pulmonary Tuberculosis. (Springer Publishing Company, 1955).

55. Ewels, P., Magnusson, M., Lundin, S. & Käller, M. MultiQC: Summarize analysis results for multiple tools and samples in a single report. Bioinformatics 32, 3047– 3048 (2016).

56. Bolger, A. M., Lohse, M. & Usadel, B. Trimmomatic: A flexible trimmer for Illumina sequence data. Bioinformatics 30, 2114–2120 (2014).

57. Li, H. et al. The Sequence Alignment/Map format and SAMtools. Bioinformatics 25, (2009).

58. Love, M. I., Huber, W. & Anders, S. Moderated estimation of fold change and dispersion for RNA-seq data with DESeq2. Genome Biol 15, 550 (2014).

59. Hänzelmann, S., Castelo, R. & Guinney, J. GSVA: Gene set variation analysis for microarray and RNA-Seq data. BMC Bioinformatics 14, 1–15 (2013).

60. Langfelder, P. & Horvath, S. WGCNA: An R package for weighted correlation network analysis. BMC Bioinformatics 9, 1–13 (2008).

61. Liberzon, A. et al. Molecular signatures database (MSigDB) 3.0. Bioinformatics 27, 1739–1740 (2011).

62. Yu, G., Wang, L. G., Han, Y. & He, Q. Y. clusterProfiler: an R package for comparing biological themes among gene clusters. OMICS 16, 284–287 (2012).

63. Yaari, G., Bolen, C. R., Thakar, J. & Kleinstein, S. H. Quantitative set analysis for gene expression: A method to quantify gene set differential expression including gene-gene correlations. Nucleic Acids Res 41, 1–11 (2013).

64. Benjamini, Y. & Hochberg, Y. Controlling the False Discovery Rate: A Practical and Powerful Approach to Multiple Testing. Journal of the Royal Statistical Society: Series B (Methodological) 57, 289–300 (1995).

